# Determinants of the COVID-19 Vaccine Hesitancy Spectrum

**DOI:** 10.1101/2021.08.05.21261675

**Authors:** Rachael Piltch-Loeb, Diana Silver, Yeerae Kim, Hope Norris, Elizabeth McNeill, David Abramson

**Affiliations:** School of Global Public Health, New York University, New York, NY 10003, USA

## Abstract

Polls report nearly one-third of the United States population is skeptical or opposed to getting the COVID-19 vaccine. Most of these polls, as well as the scientific research that has been conducted on vaccine hesitancy, was done prior to vaccine eligibility opening to all adults. Now that COVID-19 vaccines are widely available, further research is needed to understand the factors contributing to vaccine intentions across the vaccine hesitancy spectrum. This study conducted an online survey using the Social Science Research Solution (SSRS) Opinion Panel web panelists, representative of U.S. adults age 18 and older who use the internet, with an oversample of rural-dwelling and minority populations between April 8 and April 22, 2021- as vaccine eligibility opened to the country. We examined the relationship between COVID-19 exposure and socio-demographics with vaccine intentions [eager-to-take, wait-and-see, undecided, refuse] among the unvaccinated using multinomial logistic regressions [ref: fully/partially vaccinated]. Results showed vaccine intentions varied by demographic characteristics and risk exposures during the period that eligibility for the vaccine was extended to all adults.

**Funding statement:** Funding for this research was provided by a grant from the National Science Foundation (Grant #2049886). The funders had no role in study design, data collection and analysis, decision to publish, or preparation of the manuscript.

## Introduction

COVID-19 vaccines are widely available in the United States, but large portions of the population remain unvaccinated. Weekly vaccinations peaked in April 2021 as vaccine eligibility opened to all adults, and within three months had plateaued at just over half the population greater than 12 years old (1). According to a study of 39 nationally representative polls taken in the beginning of 2021, approximately one third of the population appears skeptical or unwilling to take the vaccine, and as of July 2021 vaccination rates in some states are below 40% (2). Most of these polls, as well as the scientific research that has been conducted on vaccine hesitancy, was conducted prior to this pivotal period in April when vaccine supply became available to everyone. Furthermore, the public evaluation of vaccine safety and efficacy should be considered within the empirical context of over 170 million individuals who have been fully vaccinated, the rarity of adverse events, and the apparent effectiveness of the vaccines at preventing COVID-19 hospitalizations and death. The persistence of substantial vaccine reluctance despite the overwhelming evidence of its utility and the ready accessibility to vaccine stocks suggests that there may be cultural, historical, and political cues that shape people’s vaccine intentions. Understanding the factors that contribute to such reluctance is essential to designing appropriate risk communication strategies that promote effective protective behaviors. This is particularly critical given a public health threat that has both individual and population repercussions.

A growing body of literature has determined that vaccine hesitancy is a complex decision-making process influenced by experience, risk perception, culture, confidence in authorities and medicine (3, 4). Vaccine hesitancy exists on a continuum ranging from overt acceptance to uncertainty, delay, and outright refusal. Several scholars have also found that movement along this continuum of hesitancy can be a dynamic process, with individuals’ views changing in relation to available and trusted information, social context, and personal beliefs (3). Prior to the COVID-19 vaccine rollout, several scholars investigated the relationship among vaccine hesitancy and economic and demographic factors using traditional surveys or online panels (5-11). While most studies found that Black adults were more likely to be vaccine hesitant than white adults, findings regarding other demographic characteristics were more mixed (2, 12-15). For instance, Viswanath found that those who were younger were more likely to be vaccine hesitant than those who were older, while Karpman found that those 35-49 were more likely to be vaccine hesitant, compared to both older and younger groups (12, 15). Findings regarding gender were similarly mixed, with several studies reporting that women were more likely to be vaccine hesitant, others finding the opposite, and some finding no relationship (12, 13). Findings regarding employment status and education status were similarly mixed (13-15). Few studies examined religion, but Oglagoke (14) found that vaccine hesitancy was associated with greater religiosity, although the relationship was partially mediated by respondents’ perceptions of locus of control.

In addition to demographics, some authors have explored the role of risk perception and exposure as it relates to COVID-19 vaccination intentions. For instance, Viswanath and colleagues examined the relationship between intent to vaccinate and the construct of risk perception, operationalized as susceptibility and severity (15). While higher perceptions of both susceptibility and severity were associated with intentions to take the vaccine, having someone in the family who contracted COVID-19 was not associated with intent to vaccinate. At the same time, few studies have examined the role of economic impacts of the COVID-19 pandemic on vaccine hesitancy.

The current research on COVID-19 vaccine intentions is mixed in both measurement of the hesitancy outcome and demographic results. Many research studies and polling on COVID-19 vaccination intentions have not fully captured the spectrum of intent, and focused instead on dichotomous outcomes (yes/no), 4 -point Likert scales to capture likelihood in intent, or used an outcome that has three categories (intends to be vaccinated/unsure/does not intend to be vaccinated) (2). Allowing for a wider range of responses along a continuum of intent/action may provide important insights for increasing vaccination rates. More importantly, little research has focused on the period *after* vaccines became available and plentiful and while the public health community was mobilized to promote the vaccine. This study is positioned to address these gaps using results from a nationally representative online survey in April 2021 to examine demographic and risk perception predictors of the full spectrum of vaccine hesitancy.

## Methods

### Study design and data collection

Here we describe our survey, consistent with the Checklist for Reporting Results of Internet E-Surveys (CHERRIES) (16). An online survey was conducted from a random sample of the Social Science Research Solution (SSRS) Opinion Panel web panelists, representative of U.S. adults age 18 and older who use the internet. The SSRS Opinion Panel uses two sampling methods: 1) an Address-Based Sample (ABS) frame to randomly recruit nationally representative samples, and 2) the SSRS Omnibus survey, a multi-frame random digit dial sample of landlines and cellphones, to recruit harder-to-reach demographic groups. For this study, the SSRS Omnibus survey platform oversampled Blacks, Hispanics, and adults living in rural areas. Survey items were pilot tested with unvaccinated graduate students for cognitive testing prior to administration. The 30-minute pre-tested web survey was emailed to the web panelists with a unique link with a passcode either in English or Spanish based on the respondent’s preferred language. As an incentive for participation, a gift card in electronic form was emailed after the survey. To ensure the proper administration of the survey and the content of the questionnaire, a “soft launch” was conducted on April 7, 2021, to a limited number of the respondents. The survey was fully launched to the rest of the sample on April 8, 2021 and completed on April 22, 2021. SSRS used “trap” questions as standard quality control checks in their survey and the respondents who answered three or more questions incorrectly were removed from the analysis sample. Those whose survey length was less than 5.5 minutes (less than 20% of the average length for the full sample) as well as those who skipped more than 10% of the questions were eliminated from the sample. The respondents who did not answer their vaccination status (which was the outcome variable) were also eliminated. Out of a total 6,233 samples invited to participate, we were left with 3013 respondents as our final analytic sample. IRB approval was obtained through the NYU UCAIHS and the panelists were provided with a consent form at the beginning of the survey, which they acknowledged by clicking yes and proceeding to the survey.

### Outcome variable

Survey respondents were asked whether they have received the COVID-19 vaccine. The response options were 1) fully vaccinated 2) partially vaccinated 3) not yet vaccinated. Those who had not yet been vaccinated were asked about their likelihood of taking the COVID-19 vaccine once becoming eligible. To capture the spectrum of vaccine hesitancy, answer categories included: 1) take it as soon as possible (eager), 2) wait to see how it goes before taking it (wait-and-see), 3) undecided, and 4) will not take the vaccine (refuse). Fully or partially vaccinated respondents were combined into a single category, resulting in five categories for our dependent variable.

### Independent variables

The independent variables included sociodemographic characteristics as well as COVID-19 exposure variables. For the sociodemographic characteristics, age (18-29, 30-49, 50-64 and 65 and older), sex (female and male), race/ethnicity (non-Hispanic White, non-Hispanic Black, Hispanic and other), educational attainment (Less than or graduated High School, some college or graduated college, and post graduate or professional degree), annual household income (≤$25,000, $25,001-≤$50,000, $50,001-≤$75,000, $75,001-≤$100,000, ≥$100,001), religion (Protestant, Evangelical Catholic, other and Agnostic/Atheist), living in rural or metro, types of health insurance (private, Medicare, Medicaid, TRICARE/Indian HS/Veteran/Other and uninsured), being a parent (those living with children under age 18), and political party affiliation (Democrat, Republican, independent, and don’t know) were included. As COVID-19 risk perception variables, we asked whether the respondent had been infected with COVID-19, personally knew someone who died of COVID-19, or was financially impacted by COVID-19 in the form of losing a job, losing income, and/or trouble paying for rent and other necessities.

### Statistical analysis

Descriptive and bivariate analyses used chi-square tests to evaluate associations between the distributions of the sociodemographic characteristics as well as the COVID-19 experiences with the five different outcome categories. Chi-square tests of the fully/partially vaccinated vs. take it as soon as possible group, and the wait-and-see vs. undecided group revealed that the groups were statistically significantly different from each other, justifying the use of the five categories for our study. We used unadjusted multinomial logistic regression models to estimate the independent effect of each predictor, then built an adjusted multinomial logistic regression model with all the covariates. We also tested a series of models with interaction terms (political party and age, education and race, gender and age/education/race, ethnicity and knowing someone who died of COVID personally, and education and financial hardship). Although we considered using ordered logistic regression, the data did not meet the proportional odds assumption, nor would such an approach allow us to maintain the fully/partially vaccinated group as a consistent reference group. A forest plot was drawn after running the multivariable multinomial regression to assess the effect size of each variable. A multicollinearity test was conducted by examining the conditionings of the indices (using Stata “coldiag2”) and a condition number of 30 or greater was used as a cut-off value (17). A Generalized Hosmer-Lemeshow goodness of fit test was used to assess the model fit. Data analysis was performed using Stata version 15 and a p-value of 0.05 was used as a threshold for statistical significance.

## Results

### Sample characteristics

Table 1 shows the distribution of sociodemographic characteristics as well as the COVID-19 exposure variables by the vaccine acceptance groups. In our sample, more than half of the respondents [59%] were either fully or partially vaccinated at the time of our survey. About 10% of the respondents reported that they were eager to be vaccinated, while the rest of the respondents’ [approx. 30%] answers fell along the COVID-19 vaccine hesitancy spectrum. The largest age stratum within the sample was between 30-49 years of age [37%], 55% were male, 56.4% were white non-Hispanics, either with some college education or were college graduates [51.5%], currently employed [64.4%] with private health insurance [43.8%], living in metropolitan areas [82.1%], and did not have children under the age of 18 [70.4%]. Over 85% of the sample had not had COVID-19, and the majority had not known anyone personally who died of COVID-19 [59.2%]. Over 40% of the respondents indicated that they had experienced financial hardships resulting from the pandemic. Chi-squared tests show that the distributions of each variable are statistically significantly different by the vaccine acceptance groups.

**Table 1.** Sociodemographic characteristics and COVID risk perception of the samples by vaccination status

|  |  | All [n=3013] | Fully/partially vaccinated [n=1779, 59.0%] | Eager-to-take [n= 313, 10.4%] | Wait-and-see [n=274, 9.1%] | Undecided [n=310, 10.3%] | Refuse [n=337, 11.2%] | p-value |
| --- | --- | --- | --- | --- | --- | --- | --- | --- |
| **Age groups** |  |  |  |  |  |  |  | <0.001 |
|  | 18-29 | 474 [15.8%] | 205 [11.6%] | 84 [27.0%] | 73 [26.7%] | 56 [18.2%] | 56 [16.7%] |  |
|  | 30-49 | 1112 [37.1%] | 527 [29.8%] | 144 [46.3%] | 134 [49.1%] | 146 [47.4%] | 161 [47.9] |  |
|  | 50-64 | 786 [26.2%] | 508 [28.7%] | 66 [21.2%] | 49 [18.0%] | 82 [26.6%] | 81 [24.1%] |  |
|  | 65+ | 625 [20.9%] | 529 [29.9%] | 17 [5.5%] | 17 [6.2%] | 24 [7.8%] | 38 [11.3%] |  |
| **Gender** |  |  |  |  |  |  |  | <0.001 |
|  | Female | 1370 [45.7%] | 845 [47.8%] | 163 [52.8%] | 119 [43.8%] | 122 [39.4%] | 121 [36.0%] |  |
|  | Male | 1626 [54.3%] | 924 [52.2%] | 146 [47.3%] | 153 [56.3%] | 188 [60.7%] | 215 [64.0%] |  |
| **Race/Ethnicity** |  |  |  |  |  |  |  | <0.001 |
|  | White Non-Hispanic | 1684 [56.4%] | 1053 [59.7%] | 154 [49.5%] | 123 [45.2%] | 163 [53.3%] | 191 [57.0%] |  |
|  | Black Non-Hispanic | 569 [19.0%] | 313 [17.7%] | 41 [13.2%] | 74 [27.2%] | 68 [22.2%] | 73 [21.8%] |  |
|  | Hispanic | 531 [17.8%] | 260 [14.7%] | 77 [24.8%] | 64 [23.5%] | 67 [21.9%] | 63 [18.8%] |  |
|  | Other | 204 [6.8%] | 138 [7.8%] | 39 [12.5%] | 11 [4.0%] | 8 [2.6%] | 8 [2.4%] |  |
| **Education** |  |  |  |  |  |  |  | <0.001 |
|  | Less than/graduated high school | 614 [20.4%] | 256 [14.4%] | 85 [27.2%] | 62 [22.6%] | 100 [32.4%] | 111 [32.9%] |  |
|  | Some college or graduated college | 1551 [51.5%] | 885 [49.8%] | 151 [48.2%] | 165 [60.2%] | 171 [55.3%] | 179 [53.1%] |  |
|  | Post-graduate/professional | 847 [28.1%] | 638 [35.9%] | 77 [24.6%] | 47 [17.2%] | 38 [12.3%] | 47 [14.0%] |  |
| **Employment status** |  |  |  |  |  |  |  | 0.045 |
|  | Unemployed | 1071 [35.6%] | 663 [37.3%] | 116 [37.1%] | 85 [31.0%] | 93 [30.0%] | 114 [33.8%] |  |
|  | Employed (full or part time) | 1940 [64.4%] | 1114 [62.7%] | 197 [62.9%] | 189 [69.0%] | 217 [70.0%] | 223 [66.2%] |  |
| **Annual Income** |  |  |  |  |  |  |  | <0.001 |
| | Less than \$25,000 | 515 [17.1%] | 207 [11.6%] | 68 [21.8%] | 69 [25.3%] | 78 [25.2%] | 93 [27.6%] | |
| | \$25,000 to less than \$50,000 | 656 [21.8%] | 348 [19.6%] | 80 [25.6%] | 69 [25.3%] | 83 [26.8%] | 76 [22.6%] | |
| | \$50,000 to less than \$75,000 | 573 [19.0%] | 361 [20.3%] | 48 [15.4%] | 46 [16.9%] | 62 [20.0%] | 56 [16.6%] | |
| | \$75,000 to less than \$100,000 | 464 [15.4%] | 294 [16.5%] | 38 [12.2%] | 36 [13.2%] | 49 [15.8%] | 47 [14.0%] | |
| | \$100,000 or more | 803 [26.7%] | 569 [32.0%] | 78 [25.0%] | 53 [19.4%] | 38 [12.3%] | 65 [19.3%] | |
| **Religion** |  |  |  |  |  |  |  | <0.001 |
|  | Protestant | 650 [21.6%] | 368 [20.8%] | 45 [14.4%] | 71 [26.0%] | 75 [24.2%] | 91 [27.1%] |  |
|  | Evangelical | 193 [6.4%] | 91 [5.1%] | 13 [4.2%] | 28 [10.3%] | 29 [9.4%] | 32 [9.5%] |  |
|  | Catholic, Roman Catholic | 633 [21.1%] | 399 [22.5%] | 74 [23.6%] | 41 [15.0%] | 64 [20.7%] | 55 [16.4%] |  |
|  | Other | 635 [21.1%] | 398 [22.5%] | 52 [16.6%] | 52 [19.1%] | 59 [19.0%] | 74 [22.0%] |  |
|  | Nothing in particular/Atheist/Agnostic | 894 [29.8%] | 517 [29.2%] | 129 [41.2%] | 81 [29.7%] | 83 [26.8%] | 84 [25.0%] |  |
| **Metro Status** |  |  |  |  |  |  |  | <0.001 |
|  | Non-metro | 534 [17.9%] | 304 [17.3%] | 31 [10.0%] | 41 [15.4%] | 64 [20.9%] | 94 [28.2%] |  |
|  | Metro | 2444 [82.1%] | 1456 [82.7%] | 280 [90.0%] | 226 [84.6%] | 243 [79.2%] | 239 [71.8%] |  |
| **Census region** |  |  |  |  |  |  |  | <0.001 |
|  | North East | 546 [18.3%] | 335 [19.0%] | 80 [25.7%] | 45 [16.7%] | 43 [14.0%] | 43 [12.8%] |  |
|  | North Central | 625 [20.9%] | 366 [20.7%] | 62 [19.9%] | 58 [21.6%] | 60 [19.5%] | 79 [23.6%] |  |
|  | South | 1158 [38.7%] | 664 [37.6%] | 96 [30.9%] | 121 [45.0%] | 132 [42.9%] | 145 [43.3%] |  |
|  | West | 661 [22.1%] | 402 [22.8%] | 73 [23.5%] | 45 [16.7%] | 73 [23.7%] | 68 [20.3%] |  |
| **Health Insurance** |  |  |  |  |  |  |  | <0.001 |
|  | Private | 1566 [43.8%] | 946 [53.2%] | 173 [55.3%] | 144 [52.6%] | 159 [51.3%] | 144 [42.7%] |  |
|  | Medicare | 666 [24.1%] | 530 [29.8%] | 24 [7.7%] | 28 [10.2%] | 41 [13.2%] | 43 [12.8%] |  |
|  | Medicaid | 400 [17.8%] | 156 [8.8%] | 60 [19.2%] | 56 [20.4%] | 53 [17.1%] | 75 [22.3%] |  |
|  | TRICARE/VA/Indian Health Service/Other | 187 [6.2%] | 101 [5.7%] | 20 [6.4%] | 18 [6.6%] | 21 [6.8%] | 27 [8.0%] |  |
|  | Uninsured | 193 [6.4%] | 45 [2.5%] | 36 [11.5%] | 28 [10.2%] | 36 [11.6%] | 48 [14.2%] |  |
| **Parent** |  |  |  |  |  |  |  | <0.001 |
|  | No | 2109 [70.4%] | 1375 [77.6%] | 205 [65.7%] | 163 [60.4%] | 189 [62.0%] | 177 [52.7%] |  |
|  | Yes | 886 [29.6%] | 397 [22.4%] | 107 [34.3%] | 107 [39.6%] | 116 [38.0%] | 159 [47.3%] |  |
| **Political Party** |  |  |  |  |  |  |  | <0.001 |
|  | Republican | 713 [23.7%] | 352 [19.8%] | 41 [13.1%] | 78 [28.5%] | 106 [34.2%] | 136 [40.4%] |  |
|  | Democrat | 1162 [38.6%] | 832 [46.8%] | 124 [39.6%] | 75 [27.4%] | 74 [23.9%] | 57 [16.9%] |  |
|  | Independent | 996 [33.1%] | 528 [29.7%] | 133 [42.5%] | 106 [38.7%] | 111 [35.8%] | 118 [35.0%] |  |
|  | Other | 142 [4.7%] | 67 [3.8%] | 15 [4.8%] | 15 [5.5%] | 19 [6.1%] | 26 [7.7%] |  |
| **Have you had COVID-19** |  |  |  |  |  |  |  | <0.001 |
|  | No | 2606 [86.5%] | 1580 [88.8%] | 265 [84.7%] | 232 4.7%] | 259 [83.6%] | 270 [80.1%] |  |
|  | Yes | 407 [13.5%] | 199 [11.2%] | 48 [15.3%] | 42 [15.3%] | 51 [16.5%] | 67 [19.9%] |  |
| **Personally know anyone who died of COVID-19** |  |  |  |  |  |  |  | <0.001 |
|  | No | 1785 [59.2%] | 981 [55.1%] | 199 [63.6%] | 178 [65.0%] | 195 [62.9%] | 232 [68.8%] |  |
|  | Yes | 1228 [40.8%] | 798 [44.9%] | 114 [36.4%] | 96 [35.0%] | 115 [37.1%] | 105 [31.2%] |  |
| **Financial severity\*** |  |  |  |  |  |  |  | <0.001 |
|  | 0 | 1860 [61.7%] | 1230 [69.1%] | 172 [55.0%] | 139 [50.7%] | 153 [49.5%] | 166 [49.3%] |  |

|  | All [n=3013] | Fully/partially vaccinated<br>[n=1779, 59.0%] | Eager-to-take<br>[n= 313, 10.4%] | Wait-and-see<br>[n=274, 9.1%] | Undecided<br>[n=310, 10.3%] | Refuse<br>[n=337, 11.2%] | p-value |
| --- | --- | --- | --- | --- | --- | --- | --- |
| 1 | 844 [28.0%] | 422 [23.7%] | 105 [33.6%] | 85 [31.0%] | 107 [34.5%] | 125 [37.1%] |  |
| 2 | 186 [6.2%] | 83 [4.7%] | 27 [8.6%] | 30 [11.0%] | 22 [7.1%] | 24 [7.1%] |  |
| 3 | 123 [4.1%] | 44 [2.5%] | 9 [2.9%] | 20 [7.3%] | 28 [9.0%] | 22 [6.5%] |  |

\* Financial severity is an index score for losing job, losing income, and trouble paying rent or other necessities in a scale of 0 to 3. 0 indicates no financial difficulty and 3 indicates financial difficulties in all 3 areas.

### Multinomial logistic regressions

We conducted both unadjusted and adjusted multivariable multinomial regressions to estimate the independent and controlled effect of each independent variable on vaccine acceptance. Results of unadjusted multinomial regressions are shown in Table 2. Below we describe the results of adjusted multinomial regression for each level of the vaccine hesitancy spectrum compared to those that were fully or partially vaccinated as a reference group (Table 3). Effect size and range of relative risk ratios can be seen in Figure 1.

**Table 2.** Unadjusted multinomial regression of unvaccinated groups compared to fully/partially vaccinated group as a reference category

|  |  | Relative risk of each unvaccinated group compared to fully/partially vaccinated group [RRR, 95%CI] |  |  |  |
| --- | --- | --- | --- | --- | --- |
|  |  | Eager-to-take | Wait-and-see | Undecided | Refuse |
| **Demographic characteristics** |  |  |  |  |  |
| **Age groups** |  |  |  |  |  |
|  | 18-29 | 12.75 [7.39, 22.00] * | 11.08 [6.38, 19.24] * | 6.02 [3.64, 9.97] * | 3.80 [2.44, 5.92] * |
|  | 30-49 | 8.50 [5.07, 14.26] * | 7.91 [4.71, 13.29] * | 6.11 [3.90, 9.56] * | 4.25 [2.93, 6.18] * |
|  | 50-64 | 4.04 [2.34, 6.98] * | 3.00 [1.71, 5.28] * | 3.56 [2.22, 5.70] * | 2.22 [1.48, 3.33] * |
|  | 65+ | - | - | - | - |
| **Gender** |  |  |  |  |  |
|  | Female | - | - | - | - |
|  | Male | 0.82 [0.64, 1.04] | 1.18 [0.91, 1.52] | 1.41 [1.10, 1.80] * | 1.62 [1.28, 2.07] * |
| **Race/Ethnicity** |  |  |  |  |  |
|  | White Non-Hispanic | - | - | - | - |
|  | Black Non-Hispanic | 0.90 [0.62, 1.29] | 2.02 [1.48, 2.77] * | 1.40 [1.03, 1.91] * | 1.29 [0.95, 1.73] |
|  | Hispanic | 2.02 [1.49, 2.75] * | 2.11 [1.51, 2.93] * | 1.66 [1.21, 2.28] * | 1.34 [0.97, 1.83] |
|  | Other | 1.93 [1.30, 2.86] * | 0.68 [0.36, 1.30] | 0.37 [0.18, 0.78] * | 0.32 [0.15, 0.66] * |
| **Education** |  |  |  |  |  |
|  | Less than/graduated high school | 2.75 [1.96, 3.87] * | 3.29 [2.19, 4.93] * | 6.56 [3.49, 9.79] * | 5.89 [4.06, 8.53] * |
|  | Some college or graduated college | 1.41 [1.05, 1.89] * | 2.53 [1.80, 3.55] * | 3.24 [2.25, 4.68] * | 2.75 [1.96, 3.85] * |
|  | Post-graduate/professional | - | - | - | - |
| **Employment status** |  |  |  |  |  |
|  | Unemployed | 0.99 [0.77, 1.27] | 0.76 [0.57, 0.99] * | 0.72 [0.55, 0.94] * | 0.86 [0.67, 1.10] |
|  | Employed (full or part time) | - | - | - | - |
| **Annual Income** |  |  |  |  |  |
| | Less than \$25,000 | 2.40 [1.67, 3.44] * | 3.58 [2.42, 5.29] * | 5.64 [3.71, 8.58] * | 3.93 [2.76, 5.61] * |
| | \$25,000 to less than \$50,000 | 1.68 [1.19, 2.35] * | 2.13 [1.45, 3.12] * | 3.57 [2.38, 5.36] * | 1.91 [1.33, 2.73] * |
| | \$50,000 to less than \$75,000 | 0.97 [0.66, 1.42] | 1.37 [0.90, 2.07] | 2.57 [1.68, 3.93] * | 1.36 [0.93, 1.99] |
| | \$75,000 to less than \$100,000 | 0.94 [0.62, 1.42] | 1.31 [0.84, 2.05] | 2.50 [1.60, 3.90] * | 1.40 [0.94, 2.09] |
| | \$100,000 or more | - | - | - | - |
| **Religion** |  |  |  |  |  |
|  | Protestant | - | - | - | - |
|  | Evangelical | 1.17 [0.60, 2.26] | 1.59 [0.97, 2.61] | 1.56 [0.96, 2.54] | 1.42 [0.89, 2.26] |
|  | Catholic, Roman Catholic | 1.52 [1.02, 2.26] * | 0.53 [0.35, 0.80] * | 0.79 [0.55, 1.13] | 0.56 [0.39, 0.80] * |
|  | Other | 1.07 [0.70, 1.63] | 0.68 [0.46, 1.00] | 0.73 [0.50, 1.05] | 0.75 [0.54, 1.05] |
|  | Nothing in particular/Atheist/Agnostic | 2.04 [1.42, 2.94] * | 0.81 [0.57, 1.15] | 0.79 [0.56, 1.11] | 0.66 [0.47, 0.91] * |
| **Metro Status** |  |  |  |  |  |
|  | Non-metro/rural | - | - | - | - |
|  | Metro | 1.89 [1.28, 2.79] * | 1.15 [0.81, 1.64] | 0.79 [0.59, 1.07] | 0.53 [0.41, 0.69] * |
| **Census region** |  |  |  |  |  |
|  | North East | - | - | - | - |
|  | North Central | 0.71 [0.49, 1.02] | 1.18 [0.78, 1.79] | 1.28 [0.84, 1.94] | 1.68 [1.13, 2.51] * |
|  | South | 0.61 [0.44, 0.83] * | 1.36 [0.94, 1.96] | 1.55 [1.07, 2.24] * | 1.70 [1.18, 2.45] * |
|  | West | 0.76 [0.54, 1.08] | 0.83 [0.54, 1.29] | 1.41 [0.94, 2.12] | 1.32 [0.88, 1.98] |
| **Health insurance type** |  |  |  |  |  |
|  | Private | - | - | - | - |
|  | Medicare | 0.25 [0.16, 0.38] * | 0.35 [0.23, 0.53] * | 0.46 [0.32, 0.66] * | 0.53 [0.37, 0.76] * |
|  | Medicaid | 2.10 [1.50, 2.95] * | 2.36 [1.66, 3.35] * | 2.02 [1.42, 2.88] * | 3.16 [2.28, 4.38] * |
|  | TRICARE/ Veterans/Indian HS/ Other | 1.08 [0.65, 1.80] | 1.17 [0.69, 1.99] | 1.24 [0.75, 2.04] | 1.76 [1.11, 2.78] |
|  | Uninsured | 4.37 [2.74, 6.98] * | 4.09 [2.47, 6.76] * | 4.76 [2.98, 7.61] * | 7.01 [4.50, 10.91] * |
| **Parent** |  |  |  |  |  |
|  | No | - | - | - | - |
|  | Yes | 1.81 [1.40, 2.34] * | 2.27 [1.74, 2.97] * | 2.13 [1.64, 2.75] * | 3.11 [2.44, 3.96] * |
|  |  | Eager-to-take | Wait-and-see | Undecided | Refuse |
| Political party | *Democrat* | - | - | - | - |
|  | *Republican* | 0.78 [0.54, 1.14] | 2.46 [1.75, 3.46] * | 3.39 [2.45, 4.67] * | 5.64 [4.04, 7.87] * |
|  | *Independent* | 1.69 [1.29, 2.21] * | 2.23 [1.62, 3.05] * | 2.36 [1.73, 3.23] * | 3.26 [2.33, 4.56] * |
|  | *Other, don't know, refused* | 1.50 [0.83, 2.71] | 2.48 [1.35, 4.56] * | 3.19 [1.82, 5.59] * | 5.66 [3.34, 9.59] * |
| COVID exposure/experience |  |  |  |  |  |
| Had COVID-19 | *No* | - | - | - | - |
|  | *Yes* | 1.44 [1.02, 2.02] * | 1.44 [1.00, 2.06] | 1.56 [1.12, 2.18] * | 1.97 [1.45, 2.67] * |
| Personally know anyone died of COVID-19 | *No* | - | - | - | - |
|  | *Yes* | 0.70 [0.55, 0.90] * | 0.66 [0.51, 0.86] * | 0.72 [0.57, 0.93] * | 0.56 [0.43, 0.71] * |
| Financial severity (mean, SD) ** |  | 1.40 [1.20, 1.63] * | 1.72 [1.48, 1.99] * | 1.74 [1.51, 2.00] * | 1.64 [1.42, 1.88] * |

\* p-value <0.05
\*\* Financial severity is an index score of financial hardships including losing a job, losing income, trouble paying for necessities. The minimum is 0, indicating no financial hardship and maximum is 3 indicating hardships in all three areas.

**Table 3.** Adjusted multivariable multinomial regression of unvaccinated groups compared to fully/partially vaccinated group as a reference category (n=2899)

|  |  | Relative risk of each unvaccinated group compared to fully/partially vaccinated group [RRR, 95%CI] |  |  |  |
| --- | --- | --- | --- | --- | --- |
|  |  | Eager-to-take<br>n=302 | Wait-and-see<br>n=256 | Undecided<br>n=296 | Refuse<br>n=328 |
| Demographic characteristics |  |  |  |  |  |
| Age groups | *18-29* | 6.62 [3.05, 14.39] * | 9.01 [4.04, 20.1] * | 5.63 [2.69, 11.80] * | 2.26 [1.13, 4.53] * |
|  | *30-49* | 5.79 [2.71, 12.37] * | 7.57 [3.47, 16.5] * | 6.90 [3.42, 13.88] * | 2.66 [1.39, 5.08] * |
|  | *50-64* | 2.84 [1.35, 5.94] * | 2.63 [1.23, 5.63] * | 3.57 [1.84, 6.94] * | 1.61 [0.87, 2.96] |
|  | *65+* | - | - | - | - |
| Gender | *Female* | - | - | - | - |
|  | *Male* | 0.72 [0.55, 0.94] * | 1.01 [0.75, 1.36] | 1.15 [0.87, 1.52] * | 1.46 [1.10, 1.94] * |
| Race/Ethnicity | *White Non-Hispanic* | - | - | - | - |
|  | *Black Non-Hispanic* | 0.84 [0.54, 1.30] | 2.45 [1.61, 3.74] * | 2.18 [1.45, 3.27] * | 2.63 [1.74, 3.95] * |
|  | *Hispanic* | 1.07 [0.73, 1.57] | 1.72 [1.14, 2.60] * | 1.14 [0.76, 1.72] | 1.37 [0.91, 2.07] |
|  | *Other* | 1.29 [0.82, 2.04] | 0.58 [0.28, 1.23] | 0.49 [0.18, 0.78] | 0.48 [0.22, 1.05] |
| Education | *Less than/graduated high school* | 2.20 [1.45, 3.34] * | 2.40 [1.49, 3.88] * | 4.05 [2.54, 6.44] * | 3.54 [2.28, 5.51] * |
|  | *Some college or graduated college* | 1.11 [0.80, 1.54] | 1.75 [1.20, 2.55] * | 2.12 [1.42, 3.17] * | 1.74 [1.19, 2.53] * |
|  | *Post-graduate/professional* | - | - | - | - |
| Employment status | *Unemployed* | 1.70 [1.23, 2.35] * | 1.12 [0.78, 1.61] | 0.85 [0.61, 1.20] | 0.89 [0.64, 1.24] |
|  | *Employed (full or part time)* | - | - | - | - |
| Annual Income | *Less than \$25,000* | 1.55 [0.93, 2.56] | 1.81 [1.05, 3.12] * | 3.37 [1.95, 5.84] * | 1.58 [0.95, 2.62] |
| | *\$25,000 to less than \$50,000* | 1.50 [0.98, 2.30] | 1.45 [0.91, 2.31] | 2.71 [1.68, 4.37] * | 1.07 [0.68, 1.69] |
| | *\$50,000 to less than \$75,000* | 1.02 [0.67, 1.55] | 1.10 [0.69, 1.74] | 2.17 [1.36, 3.45] * | 0.97 [0.63, 1.50] |
| | *\$75,000 to less than \$100,000* | 1.00 [0.64, 1.56] | 1.11 [0.68, 1.82] | 2.04 [1.25, 3.32] * | 1.08 [0.69, 1.70] |
| | *\$100,000 or more* | - | - | - | - |
| Religion | *Protestant* | - | - | - | - |
|  | *Evangelical* | 0.83 [0.40, 1.69] | 1.30 [0.74, 2.26] | 1.29 [0.75, 2.24] | 1.08 [0.64, 1.83] |
|  | *Catholic, Roman Catholic* | 1.02 [0.64, 1.61] | 0.57 [0.35, 0.91] * | 0.91 [0.59, 1.41] | 0.65 [0.42, 1.01] |
|  | *Other* | 0.83 [0.52, 1.32] | 0.77 [0.49, 1.19] | 0.87 [0.57, 1.31] | 0.88 [0.59, 1.30] |
|  | *Nothing in particular/Atheist/Agnostic* | 1.31 [0.87, 2.00] | 0.93 [0.62, 1.40] | 0.98 [0.66, 1.46] | 0.91 [0.61, 1.34] |
| Metro Status | *Non-metro/rural* | - | - | - | - |
|  | *Metro* | 2.06 [1.32, 3.21] * | 1.22 [0.82, 1.83] | 0.95 [0.67, 1.35] | 0.71 [0.51, 0.98] * |
|  |  | Eager-to-take<br>n=302 | Wait-and-see<br>n=256 | Undecided<br>n=296 | Refuse<br>n=328 |
| **Census region** | *North East* | - | - | - | - |
|  | *North Central* | 0.68 [0.45, 1.01] | 0.93 [0.60, 1.47] | 0.95 [0.60, 1.50] | 1.20 [0.77, 1.88] |
|  | *South* | 0.55 [0.38, 0.79] * | 0.86 [0.57, 1.29] | 1.04 [0.69, 1.57] | 1.02 [0.68, 1.55] |
|  | *West* | 0.67 [0.45, 0.98] * | 0.59 [0.36, 0.95] * | 1.35 [0.87, 2.12] | 1.11 [0.70, 1.76] |
| **Health insurance type** | *Private* | - | - | - | - |
|  | *Medicare* | 0.48 [0.24, 0.93] * | 0.93 [0.49, 1.79] | 1.10 [0.61, 1.98] | 1.03 [0.56, 1.87] |
|  | *Medicaid* | 1.13 [0.73, 1.76] | 1.18 [0.74, 1.88] | 0.95 [0.60, 1.49] | 1.84 [1.19, 2.86] * |
|  | *TRICARE/ Veterans/Indian HS/ Other* | 0.97 [0.65, 1.66] | 0.88 [0.49, 1.55] | 0.84 [0.49, 1.46] | 1.36 [0.81, 2.30] |
|  | *Uninsured* | 2.43 [1.42, 4.17] * | 1.35 [0.73, 2.52] | 1.96 [1.13, 3.40] * | 4.20 [2.46, 7.18] * |
| **Parent** | *No* | - | - | - | - |
|  | *Yes* | 1.13 [0.82, 1.55] | 1.22 [0.88, 1.70] | 1.23 [0.89, 1.70] | 1.92 [1.40, 2.64] * |
| **Political party** | *Democrat* | - | - | - | - |
|  | *Republican* | 1.20 [0.78, 1.85] | 5.53 [3.59, 8.53] * | 6.66 [4.40, 10.07] * | 11.06 [7.21, 16.96] * |
|  | *Independent* | 1.69 [1.25, 2.29] * | 3.04 [2.12, 4.37] * | 3.02 [2.11, 4.33] * | 4.30 [2.93, 6.30] * |
|  | *Other, don't know, refused* | 1.16 [0.61, 2.22] | 2.02 [0.98, 4.15] | 3.16 [1.71, 5.82] * | 5.48 [3.05, 9.85] * |
| **COVID exposure/experience** |  |  |  |  |  |
| **Had COVID-19** | *No* | - | - | - | - |
|  | *Yes* | 1.47 [1.01, 2.14] * | 1.26 [0.84, 1.87] | 1.29 [0.89, 1.88] | 1.59 [1.11, 2.26] * |
| **Personally know anyone died of COVID-19** | *No* | - | - | - | - |
|  | *Yes* | 0.75 [0.57, 0.99] * | 0.65 [0.48, 0.87] * | 0.64 [0.49, 0.85] * | 0.49 [0.37, 0.66] * |
| **Financial severity (mean, SD) \*\*** |  | 1.01 [0.84, 1.20] | 1.23 [1.03, 1.46] * | 1.25 [1.06, 1.48] * | 1.21 [1.02, 1.44] * |

\* p-value <0.05
\*\* Financial severity is an index score of financial hardships including losing a job, losing income, trouble paying for necessities. The minimum is 0, indicating no financial hardship and maximum is 3 indicating hardships in all three areas.

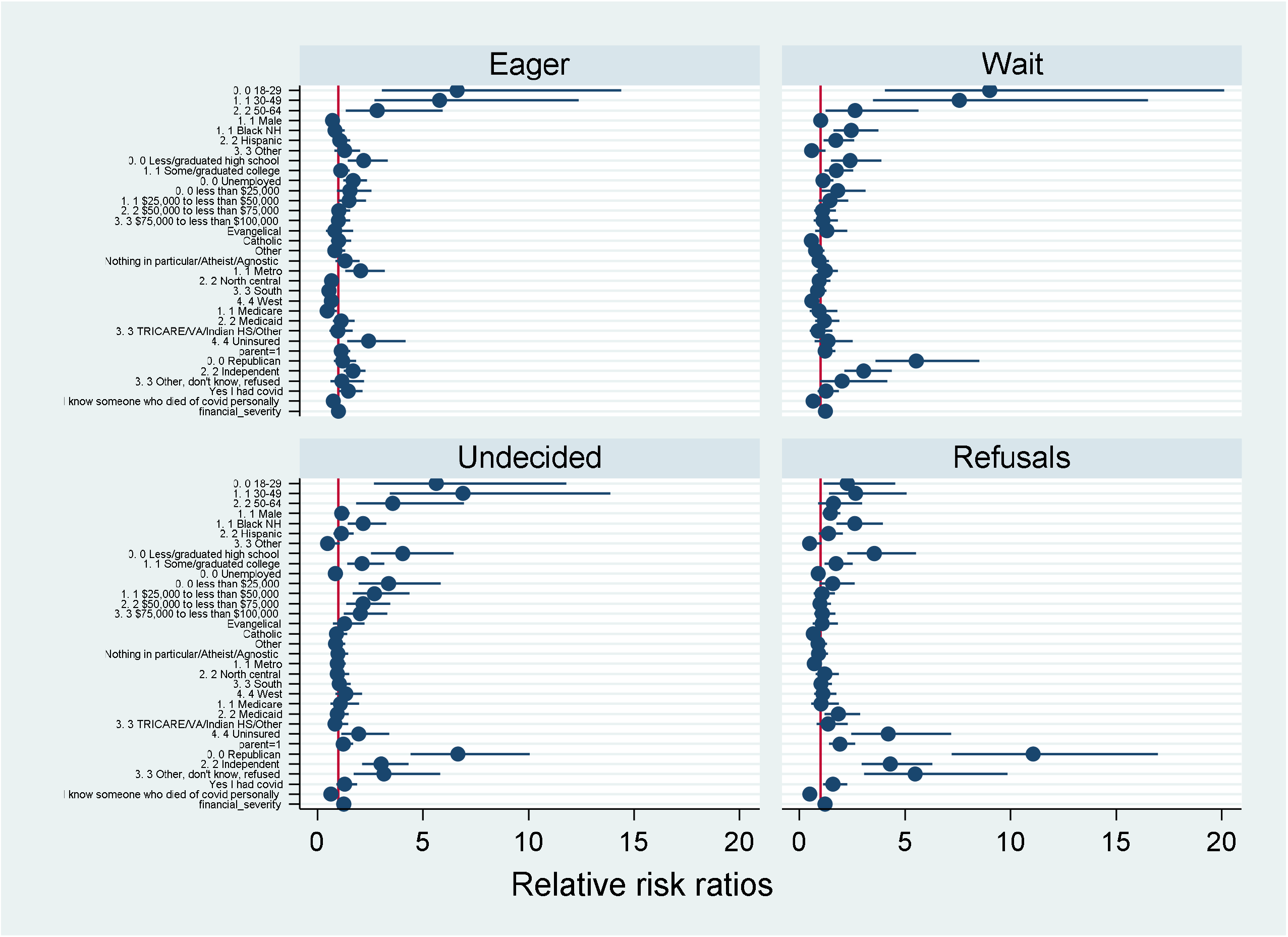

### Eager–to-take

Age was a significant predictor in the adjusted multivariable multinomial regression. Those who are younger were more likely than those over 65 to be in the eager to be vaccinated group than to be in the fully/partially vaccinated group. The relative risk ratios (RRR) of being in eager to be vaccinated group for the age groups of 18-29, 30-49 and 50-64 were 6.62 [95% CI: 3.05-14.4], 5.79 [95% CI: 2.71-12.4] and 2.84 [CI: 1.35-5.94], respectively. Although gender was not a significant predictor in unadjusted multinomial regression (see Table 2), male respondents had a significantly increased risk of being in the eager to be vaccinated group than female ones [RRR=0.72, 95%CI:0.55-0.94] when other demographic characteristics were controlled for. As for education, those who had attained less than high school or graduated high school were more likely than the college-educated to be in the eager to be vaccinated group [RRR=2.20, 95% CI:1.45-3.34] than to be in the fully/partially vaccinated group. Employment status showed a statistical significance, indicating that those who were unemployed were more likely than the employed to be in the eager to be vaccinated group [RRR=1.70, 95%CI:1.23-2.35] than in the fully/partially vaccinated group. The RRR for respondents who live in the city was 2.06, showing that they were more likely than non-city dwellers to be in the eager-to-take group. When compared to respondents who live in the Northeast region, those who live in the South and the West regions were less likely to be in the eager-to-take group [RRR=0.55, 95% CI:0.38-0.79 for South region, RRR=0.67, 95% CI:0.45-0.98 for North region] than to be in the fully/partially vaccinated group. Those who have Medicare as their primary health insurance were less likely than private insurance to be in the eager-to-take group [RRR=0.48, 95% CI:0.24-0.94]. In contrast, those without health insurance had a higher risk of being in the eager-to-take group [RRR=2.43, 95%CI:1.42-4.17] than to be fully/partially vaccinated group. The RRR for being in the eager-to-take group was 1.69 [95% CI: 1.25-2.29] for those with independent political affiliation, indicating that they were more likely to be in the eager-to-take group than to be in the fully/partially vaccinated group. Among the COVID-19 exposure variables, those who had COVID-19 were more likely than the uninfected to be in the eager-to-take group [RRR=1.47, 95% CI:1.01-2.14] than to be in the fully/partially vaccinated group. Those who knew someone who died of COVID-19 personally were less likely to be in the eager-to-take group [RRR= 0.75 [95%CO: 0.57-0.99] and more likely to be in the fully/partially vaccinated group.

### Wait-and-see

Those who are younger were more likely than those over 65 to be in the wait-and-see group than to be in the fully/partially vaccinated group. The youngest age group had the highest RRR of 9.01 [95% CI:4.04-20.1] of being in the wait-and-see group, followed by those in the age group 30-49 [RRR= 7.57, 95% CI:3.47-16.5] and 50-64 [RRR=2.64, 95% CI: 1.23-5.63]. Non-Hispanic black and Hispanic respondents were more likely than non-Hispanic white respondents to be in the wait-and-see group [RRR=2.45, 95% CI: for black non-Hispanic, RRR=1.72 for Hispanic] than to be in the fully/partially vaccinated group. Level of education continued to be a significant factor, showing that those with less education were more likely than those with post-graduate or professional degrees to be in the wait-and-see group [RRR=2.40, 95%CI: 1.49-3.88 for high school graduates or less, RRR=1.75, 95%CI: 1.20-2.55 for college graduates or less] than to be in the fully/partially vaccinated group. Compared to those in the highest income group, those who belonged to the lowest income group were more likely to be in the wait-and-see group [RRR=1.81, 95%CI: 1.05-3.12] than to be in the fully/partially vaccinated group. The respondents who are Catholics had a lower risk than those who are protestant of being in the wait-and-see group [RRR=0.57, 95% CI: 0.35-0.91]. Respondents who identified their political affiliation as Republican or Independent were more likely than Democrats to be in the wait-and-see group [RRR=6.66, 95% CI:4.40-10.1 for Republicans, RRR=3.16, 95% CI: 1.71-5.82] than to be in the fully/partially vaccinated group. Those who personally knew someone died of COVID-19 were less likely than those who did not personally know someone died of COVID-19 to be in the wait-and-see group [RRR= 0.65, 95%CI: 0.48-0.87], than to be in the fully/partially vaccinated group. Further, the more financial severity one experienced, the more likely it is for that individual to be in the wait-and-see group rather than fully/partially vaccinated [RRR=1.23, 95%CI: 1.06-1.48] group.

### Undecided

Compared to 65 and older respondents, those who were younger were more likely to be undecided than fully/partially vaccinated [RRR=5.63, 95% CI:2.69-11.8 for age group 18-29, RRR=6.90 95% CI:3.42-13.9 for age group 30-49, RRR=3.57, 95% CI: 1.84-6.94 for age group 50-64]. Compared to non-Hispanic whites, non-Hispanic black respondents had a higher risk of being undecided [RRR=2.18, 95% CI: 1.45-3.27] than fully/partially vaccinated. Those who had less education were more likely than those who had post-graduate or professional degrees to be undecided than vaccinated [RRR=4.05, 95% CI: 2.54-6.44 for high school graduates or less, RRR=2.12, 95% CI: 1.42-3.17]. Similarly, those in lower-income level groups had a higher risk than those with the highest income group of being in the undecided group [RRRs 3.17-2.04] than to be in the fully/partially vaccinated group. Respondents without health insurance had a higher risk [RRR=1.96, 95% CI:1.13-3.40] than the respondents with private health insurance to be undecided rather than fully/partially vaccinated. Compared to Democrats, all the other groups i.e., Republican, Independent, and other/don’t know/refused respondents were more likely to be in the undecided group than to be in the fully/partially vaccinated group and that risk was highest among the Republicans [RRR=7.77, 95% CI:4.40-10.1]. In terms of COVID-19 exposure, if respondents personally knew someone who died of COVID-19 compared to who did not know someone died of COVID-19, they were less likely [RRR=0.64, 95% CI: 0.49-0.85] to be undecided and more likely to be fully/partially vaccinated. In contrast, those with severe financial hardship were more likely to be undecided [RRR=1.25, 95% CI: 1.06-1.48] and less likely to be fully/partially vaccinated.

### Refusal

The respondents in the two youngest age groups were more likely than those over 65 to refuse the vaccine [RRR=2.26, 95% CI: 1.13-4.52 for age group 18-29, RRR=2.66, 95% CI: 1.39-5.08 for age group 30-49]. The RRR for males and non-Hispanic blacks were 1.46 [95%CI: 1.10-1.94] and 2.63 [95% CI:1.74-3.95] respectively, indicating that they had a higher risk than females and non-Hispanic whites to refuse the vaccine compared to fully/partially vaccinated. Following the same trend, those with less education had a higher risk than those with post-graduate or professional degrees to be in the refusal group [RRR=3.54, 95% CI:2.28-5.51 for high school graduates or less, RRR=1.74, 95% CI: 1.19-2.53] than to be in fully/partially vaccinated group. Respondents living in urban areas were less likely than those living in rural areas to be refusers [RRR=0.71, 95% CI: 0.51-0.98] and more likely to be fully/partially vaccinated. Those who have Medicaid or were uninsured were more likely than those with private health insurance to refuse to be vaccinated [RRR=1.84, 95% CI: 1.19-2.86 for Medicaid, RRR=4.20, 95% CI: 2.46-7.18 for uninsured] than fully/partially vaccinated. Parents were more likely than non-parents to be refusers [RRR=1.92, 95% CI: 1.40-2.64] than fully/partially vaccinated. Compared to Democrats, all other political affiliations were more likely to refuse the vaccine than fully/partially vaccinated, republicans showing the highest risk [RRR=11.06, 95% CI: 7.21-17.0]. All of the COVID-19 exposure variables were statistically significant factors in predicting the risk of being in the refusal groups. Those who have had COVID-19 infection and those with a severe financial hardship were more likely than those who did not have COVID-19 infection and financial hardship to refuse the vaccine [RRR=1.59, 95% CI: 1.11-2.26 for having had COVID-19, RRR=1.21, 95% CI: 1.02-1.44 for financial hardship] while those who personally knew someone who died of COVID-19 were less likely than those who did not know someone who died of COVID-19 to be refusers [RRR=0.49, 95% CI: 0.37-0.66].

The multivariable multinomial regression models with interactions terms between political party and age, education and race, gender and age, gender and education, gender and race, ethnicity and knowing someone who died of COVID-19, as well as education and severity in financial hardship, were not statistically significant.

## Discussion

As vaccination opened to the adult public, approximately 40% of our sample was unvaccinated. Those who were unvaccinated fell across four categories-eager to take the vaccine, waiting to see how it goes before deciding to take the vaccine, undecided, and refusing to take the vaccine. The proportion of our sample that was vaccinated was similar to representative national surveys conducted at similar points in time.

Our results confirm that demographic characteristics and risk exposures varied along the vaccine hesitancy spectrum during the period that eligibility for the vaccine was extended to all adults.

Unsurprisingly, younger ages predicted all categories along the spectrum, since those who were already vaccinated were more likely to be older, as part of the early eligibility for vaccines pursued by the states. As visualized in Figure 1, age had a large effect on being eager to take the vaccine or waiting to see, possibly because for younger age groups eligibility was more recent, and there were evolving social norms among peers.

Similarly, those with less educational attainment were more likely than those with a college education or more to be in one of the unvaccinated groups, compared to those who were at least partially vaccinated, which may also reflect the early eligibility for the vaccine, given that healthcare providers who often need additional education were initially eligible. However, while men were less likely to be eager to take the vaccine and more likely to refuse it, gender was not a significant predictor of the “wait and see” or the “undecided” category, all else equal. As expected, Blacks were more likely than Whites to be in every category of the hesitancy spectrum, except being eager to get the vaccine.

Employment status was significant only for those eager to get the vaccine compared to those who were vaccinated, such that those who were unemployed were 75% more likely than those who were employed to be eager to get the vaccine – perhaps viewing vaccination as a condition for working. Interestingly, compared to those who were fully vaccinated, income was a monotonic and significant predictor for being undecided about getting the vaccine. Those who were in the lowest income group compared to the highest were also nearly 80% more likely to indicate that they intended to “wait and see” before making the decision to be vaccinated.

Last, as expected, those in metropolitan areas were far more likely to be eager to get the vaccine than those in rural areas, but surprisingly, urbanicity was not a predictor for any other group. Compared to those who were fully vaccinated, region of the country was only predictive of being in the eager to get the vaccine group, where compared to the Northeast, residents of every other census region had 34-46% lesser odds of being eager to get vaccinated.

Numerous reports in the media have highlighted the role of political party membership in predicting vaccine hesitancy, and our results confirm these reports. For the “wait and see” group, the undecided group, and those who would refuse the vaccine, being anything other than a Democrat was predictive of higher odds of belonging to those three groups. Only Independents had higher odds than Democrats of being eager to receive the vaccine. The magnitude of the effect size of political party on outright refusal was particularly large.

Our findings also depict the toll that the pandemic has taken: 41% of our sample knew someone who died of COVID-19, and 38% had suffered some serious financial hardship they attributed to the pandemic. Yet, while we had hypothesized that these experiences would increase the odds of wanting to get the vaccine, we also found that these experiences increased the odds of being in all other categories compared to the fully vaccinated, suggesting that this experience did not necessarily heighten one’s willingness to get the vaccine, all else equal. Similarly, having experienced severe financial hardships resulting from the vaccine was predictive of being in the wait-and-see, undecided, and refuse groups. It may be that that people experiencing these hardships may blame government for the mitigation strategies employed for the pandemic (mask-wearing, social distancing, closing bars and restaurants, etc.) and thus also be less trustful of the vaccine development process and vaccination advice promoted by government officials. Our finding that having had COVID-19 increased the odds of being both eager to get the vaccine or refusing the vaccine confirms the ways in which these experiences operate differently than public health frameworks, such as the Health Belief Model, would suggest.

Our analysis posited differences in the association between demographic characteristics and COVID-19 exposures and impact and levels of vaccine hesitancy. We interpreted agreement with the phrase “I will wait to see how it goes before taking the vaccine” (Wait-and-see) as someone who was actively engaged in monitoring reports of vaccine safety and efficacy. For those who indicated that they were “undecided” about taking the vaccine, we hypothesized that their willingness to take the vaccine would be less than those who indicated they would wait and see, and expected their reluctance to be more diffuse. Our bivariate analyses reveal differences between these two groups. However, although the results from the multinomial logit models show differences in magnitude for the association between demographics and these outcomes, the only demographic variable that distinguished these groups was Hispanic ethnicity, such that compared to Whites, Hispanics were 73% more likely to indicate that they would “wait and see” before getting the vaccine [RRR=1.73, 95% CI: 1.14-2.60]. More research is needed to determine what differences in attitudes, beliefs, and experiences related to vaccines and COVID-19 may distinguish this group.

Many of the results reported here suggest that hesitancy about the COVID-19 vaccine may be different than vaccine hesitancy for other adult vaccines. For instance, while Okoli’s study of adult vaccine hesitancy for the flu vaccine found that younger ages were more vaccine hesitant, several other studies (Schmid, Chu) did not (18-20). Similarly, findings regarding gender are similarly mixed. However, numerous studies have found that Blacks have higher odds of vaccine hesitancy than do Whites, as our study confirms here (2, 12-15).

Like all studies, this study has some limitations. Our sample differs from the adult population in several ways, as do most online survey panels. However, a number of high-quality studies have used similar approaches, whether by using the KnowledgePanel, Amazon M Turk, or other firms, and SSRS has a robust mechanism for quality checks that were embedded in our survey. Our study is cross-sectional, and thus cannot make claims about causality. Finally, our survey relies on self-report, and focuses on intentions, not behaviors, which may change. At the same time, it offers a snapshot of the US public’s intentions just as all adults became eligible for the vaccine, such that intentions could be quickly realized.

## Conclusion

Prior research has found individuals’ relative position on the hesitancy spectrum is dynamic rather than static (21). Attention to population profiles of the vaccine hesitancy spectrum can aid public health officials in targeting new strategies for increasing vaccine acceptance. While intense media attention has focused on those who have refused the vaccine, our findings suggest that a large portion of the unvaccinated public may be reached and persuaded to move from “waiting and seeing” or being undecided to becoming vaccinated. Those who are eager to take the vaccine are the first priority as vaccine access increases. Public health leaders developing messages to reach these populations can use profiles such as these in their efforts.

## Data Availability

Data is available upon request of the authors.

